# A validated model for early prediction of group A streptococcal aetiology and clinical endpoints in necrotising soft tissue infections

**DOI:** 10.1101/2024.06.05.24308478

**Authors:** Sonja Katz, Jaco Suijker, Steinar Skrede, Annebeth Meij-de Vries, Anouk Pijpe, Anna Norrby-Teglund, Laura M Palma Medina, Jan K Damås, Ole Hyldegaard, Erik Solligård, Mattias Svensson, PerAID/PerMIT/INFECT study group, Knut Anders Mosevoll, Vitor AP Martins dos Santos, Edoardo Saccenti

## Abstract

**Objectives:** To develop and externally validate machine learning models for predicting microbial aetiology and clinical endpoints, encompassing surgery, patient management, and organ support in Necrotising Soft Tissue Infections (NSTI).

**Methods:** Predictive models for the presence of Group A Streptococcus (GAS) and for five clinical endpoints (risk of amputation, size of skin defect, maximum skin defect size, length of ICU stay, and need for renal replacement therapy) were built and trained using data from the prospective, international INFECT cohort (409 patients, 2013-2017), implementing unsupervised variable selection, and comparing several algorithms. SHapley Additive exPlanations (SHAP) analysis was used to interpret the model. GAS predictive models were externally validated using data from a Dutch retrospective multicenter cohort from the same calendar period (216 patients).

**Results:** Eight variables available pre-surgery (age, diabetes, affected anatomical locations, prior surgical interventions, and creatinine and haemoglobin levels) sufficed for prediction of GAS aetiology with high discriminatory power in both the development (ROC-AUC: 0.828; 95%CI 0.763, 0.883) and validation cohort (ROC-AUC: 0.758; 95%CI 0.696, 0.821). The prediction of clinical endpoints related to surgical, patient management, and organs support aspects was unsuccessful.

**Conclusion:** An externally validated prediction model for GAS aetiology before organ support aspects was unsuccessful, having implications for targeted treatment decisions of NSTI.

## Introduction

Necrotizing soft tissue infections (NSTI) are rare yet severe bacterial infections that impact a diverse demographic population regarding age, gender, and comorbidities (1). NSTI can be caused by either a single pathogen (monomicrobial, type II NSTI), most often *Streptococcus pyogenes* (group A streptococcus; GAS), or a combination of multiple pathogens (polymicrobial, type I NSTI), each employing distinct pathogenic mechanisms (2,3). In addition to tissue damage, NSTI is characterised by systemic toxicity, often resulting in sepsis and septic shock (28-50%) (4,5). The course of the disease is rapid, and mortality rates vary from 10-29% (5–7). Long-term consequences include extensive morbidity, manifesting in functional impairments such as amputations, scarring and fatigue with significant psychosocial impacts, including fear of recurrence, post-traumatic stress, depression, and changes in social engagement (8,9).

Given the severity of NSTI, an early overview of disease characteristics predicting outcomes could significantly improve prognosis and outcomes. A major factor in deciding the appropriate treatment revolves around ascertaining whether GAS is the causative microorganism (10), thereby guiding specific antimicrobial treatment including clindamycin (11). Whereas intravenous immunoglobulin (IVIG) is not routinely recommended for the treatment of NSTI in general, data supports it may lead to improved outcomes in case of GAS based NSTI (12,13). Therefore some guidelines recommend routinely using (14) or considering IVIG (10) in case of GAS-based NSTI. Because the earliest method to identify the likelihood of GAS involvement is a gram stain on samples obtained during surgery (14), an earlier accurate, and preferably pre-surgery, identification of these patients could reduce the time until the start of IVIG within hours. Given the recent worldwide surge in the incidence of GAS (15–17), an early accurate prediction of GAS aetiology could lead to the start of targeted treatment before results of microbiological tests are available, while avoiding over-treatment.

Applying machine learning-based predictive models holds promising potential in overcoming shortcomings related to accuracy, individualization, and applicability in clinical decision-making for NSTI patients. We have recently shown (18) that such a model can accurately predict 30-day mortality. We built and trained a predictive machine learning model using just sixteen clinical parameters/variables collected within the initial 24 hours of ICU admission enabling a more precise prediction of 30-day mortality compared to commonly used clinical scoring systems, like the Simplified Acute Physiology Score (SAPS) (19) and the Sequential Organ Failure Assessment (SOFA) (20) scores used to predict mortality in intensive care units (ICU) patients.

In this study we develop and externally validate prediction models for several clinical endpoints relevant to NSTI, including likely GAS aetiology, surgical aspects (risk of amputation, size of skin defect after fist surgery, maximum skin defect size), as well as length of ICU stay, and need for organ support (need for renal replacement therapy) .

## Methods

### Study design

#### Development cohort

Subjects and data of the cohort used to develop and train the prediction models were obtained from the INFECT study (https://permedinfect.com/; registration number NCT01790698, ClinicalTrials.gov), an international, multicentre, prospective, cohort study with patients with NSTI included prospectively at five Scandinavian hospitals (Supplementary Note 1). A total of 409 patients above the age of 18 and with surgically confirmed NSTI cases were enrolled between February 2013 and June 2017. Patients’ enrollment and data collection protocols, as well as demographics, have been published previously (2,4).

#### Validation cohort

Subjects and the predictive models were obtained from a Dutch retrospective multicenter cohort (21) comprised of 216 patients admitted for acute treatment of NSTI to 11 centres between January 1, 2013, and December 31, 2017, irrespective of a subsequent ICU admissions (Supplementary Note 1). Patient characteristics have been previously described (21).

### Clinical endpoints

Six clinically relevant NSTI endpoints were selected through semi-structured interviews as previously described (18) and were used for machine learning modelling. The selected endpoints encompass different clinical aspects of NSTI. Bacterial aetiology: presence of GAS (binary). Surgical aspects: risk of amputation (binary); size of skin defect after fist surgery, % of body surface (continuous); maximum skin defect size,% of body surface (continuous). Patient management: length of ICU stay (continuous). Organ support: need for Renal Replacement Therapy (RRT) (binary). The binary endpoints were coded as 0-1 (absence-presence, no-yes). An overview on the number of patients included and label balance for each clinical outcome can be found in Supplementary Table 1.

### Data preprocessing

For our development cohort we utilised data from within the INFECT study during which over 700 clinical variables were collected, encompassing data available from hospital admission to ICU admission (2). The clinical variables were categorised depending on their availability at four time points: entry (upon hospital admission, 45 variables), pre-surgery (prior to the first surgical procedure in the referral centre, 56 variables), post-surgery (posterior to first surgical procedure and prior to ICU admission, 723 variables), baseline (BL; first 24 h of ICU admission, 762 variables). For a graphical overview detailing the time point for data collection see Supplementary Figure 1b; the number of variables in each subset can be found in Supplementary Table 2. Data cleaning, imputation, and post-processing have been implemented as previously described (18). For validation of models, we extracted the subset of variables required for prediction from the validation cohort and preprocessed the data using the same cleaning and imputation procedure as those applied to the development cohort.

### Selection of input variables for the prediction

The selection of variables relevant for the prediction of each clinical endpoint was done through unsupervised variable selection utilising the Boruta algorithm (22). To obtain robust sets of relevant variables, we implemented an iterative version. Therein, we i) created a subset of the dataset by randomly sampling 80% of patients and ii) iteratively ran Boruta for 50 times with different initialisation on the bootstrapped dataset and iii) repeated steps i) - ii) for 30 iterations. Only variables identified in more than 50% of the iterations were considered relevant and kept for further analyses.

### Machine-learning model development

A graphical overview detailing the developmental and validation process of the machine learning models can be found in Supplementary Figure 1a.

#### Model selection

For classification models involving binary endpoints (GAS aetiology, need for RRT, risk of amputation), three different predictive models based on different statistical and machine learning approaches were developed and compared: logistic regression (23), Gaussian process classifier (24), and Random Forest classifier (25). Models for regression tasks (length of ICU stay, maximum size of skin defect) included lasso regression, ridge regression, elastic nets, Gaussian process regression (24), multi-layer perceptron regression, and Random Forest regression (25).

The predictive power of each model was compared through leave-one-out cross-validation. Model hyperparameters were optimised using an exhaustive grid search, using balanced accuracy for classification and negative mean squared error for regression as scoring metric for model selection; a summary on the hyperparameters optimised can be found in Supplementary Note 2. Only the best performing models were used for internal validation with bootstrapping.

#### Model training and internal validation

Internal validation was conducted to ensure optimism-corrected performance evaluation and quantify the uncertainty associated with our developed models. This entailed repeating the model training and hyperparameter optimisation process over 1000 bootstrap samples (*n*=1000) drawn from the development cohort, enabling the calculation of robust 95% confidence intervals (95%CIs).

#### External validation

For external validation, the generalisation performance of the trained models was assessed through bootstrapping of the validation cohort (*n* = 10000).

To enable models to account for the heterogeneity of data across time, geography and facilities, we adopted the local validation scheme proposed by (26) and fine-tuned models trained on the development cohort using 30% of the validation data.

### Model performance

The quality of the performance of classification models was evaluated using several metrics: Precision, Recall, F_1_-score, balanced accuracy, Brier score, area under the receiver-operator curve (ROC-AUC), and average precision-recall score (PR). The quality of regression models was assessed using the coefficient of determination (R²) as well as the mean absolute error (MAE) and mean squared error (MSE). Quality measures are given as mean and associated 95%CI calculated over all bootstrapping iterations. AUC are given only in the case of balanced groups as this measure can give biased indication in presence of unbalanced data.

### Model explainability

SHapley Additive exPlanations (SHAP) values were used to assess relative contribution of clinical variables to the prediction/classification models (27). SHAP values were calculated for each model training/validation step since they are sensitive to model parameterization and data splits. Results are given as mean with associated 95%CI.

### Software

For all machine-learning models the implementations available in the scikit-learn Python library (version 1.4.2) were used. For variable selection, Boruta version 0.3 was used (22). SHAP analysis was conducted using the package version 0.43.0. Our source code for the GAS predictive models is available at https://github.com/sonjakatz/permit-nsti-gas.

## Results

### Prediction of GAS aetiology in NSTI

To predict GAS aetiology in NSTI as early as possible, we trained prediction models for the presence/absence of GAS using time-dependent subsets of clinical variables, namely *entry, pre-surgery, post-surgery, baseline*. We conducted variable selection to identify a minimal set of clinically relevant variables that are feasible to obtain in a clinical setting. This process aimed to optimise the model by reducing the risk of overfitting while maintaining high prediction quality. The number of selected variables ranged from 6 (*entry*) to 14 (*baseline*) as shown in Table 1.

**Table 1.** Overview of variables yielded through unsupervised variable selection. A more detailed variable description can be found in Supplementary Table 3.

| **Entry**<br>(6/45 variables) | **Pre-surgery**<br>(8/56 variables) | **Post-surgery**<br>(9/723 variables) | **Baseline**<br>(14/762 variables) |
| --- | --- | --- | --- |
| affected<sup>1</sup>: upper arm | affected<sup>1</sup>: upper arm | affected<sup>1</sup>: upper arm | affected<sup>1</sup>: upper arm |
| affected<sup>1</sup>: lower arm | affected<sup>1</sup>: lower arm | affected<sup>1</sup>: lower arm | affected<sup>1</sup>: lower arm |
| affected<sup>1</sup>: anogenital area | affected<sup>1</sup>: anogenital area | affected<sup>1</sup>: anogenital area | affected<sup>1</sup>: anogenital area |
| surgery before | surgery before | surgery before | surgery before |
| diabetes | diabetes | diabetes | diabetes |
| age | creatinine<sup>2</sup> | creatinine<sup>2</sup> | creatinine<sup>2</sup> |
|  | haemoglobin<sup>2</sup> | haemoglobin<sup>2</sup> | haemoglobin<sup>2</sup> |
|  | age | creatinine<sup>3</sup> | creatinine<sup>3</sup> |
|  |  | Anatomical site sampled | systolic BP (lowest)<sup>4</sup> |
|  |  |  | creatinine<sup>4</sup> |
|  |  |  | noradrenaline<sup>4</sup> |
|  |  |  | platelets<sup>4</sup> |
|  |  |  | lactate<sup>4</sup> |
|  |  |  | glucose<sup>4</sup> |

<sup>1</sup> at arrival at specialised hospital
<sup>2</sup> preoperative (preop): before the first surgery, which is before ICU admission
<sup>3</sup> preadmission: upon ICU admission
<sup>4</sup> baseline (BL): during the first 24 hours in the ICU

Based on these reduced sets of variables, the performance of several machine learning-based classifiers was compared, with Random Forest classifiers standing out in terms of performance across all subsets. Therefore, only results for the Random Forest prediction model will be presented. Results for other models are available in Supplementary Table 4.

Comparison of prediction performances showed distinct differences between different time-dependent subsets (Supplementary Table 5). Notably, this revealed that performances peak using information already available *pre-surgery* (ROC-AUC 0.828; 95%CI 0.763, 0.883), constituting the earliest possible time point for prediction of GAS aetiology (Figure 1a).

**Figure 1.**
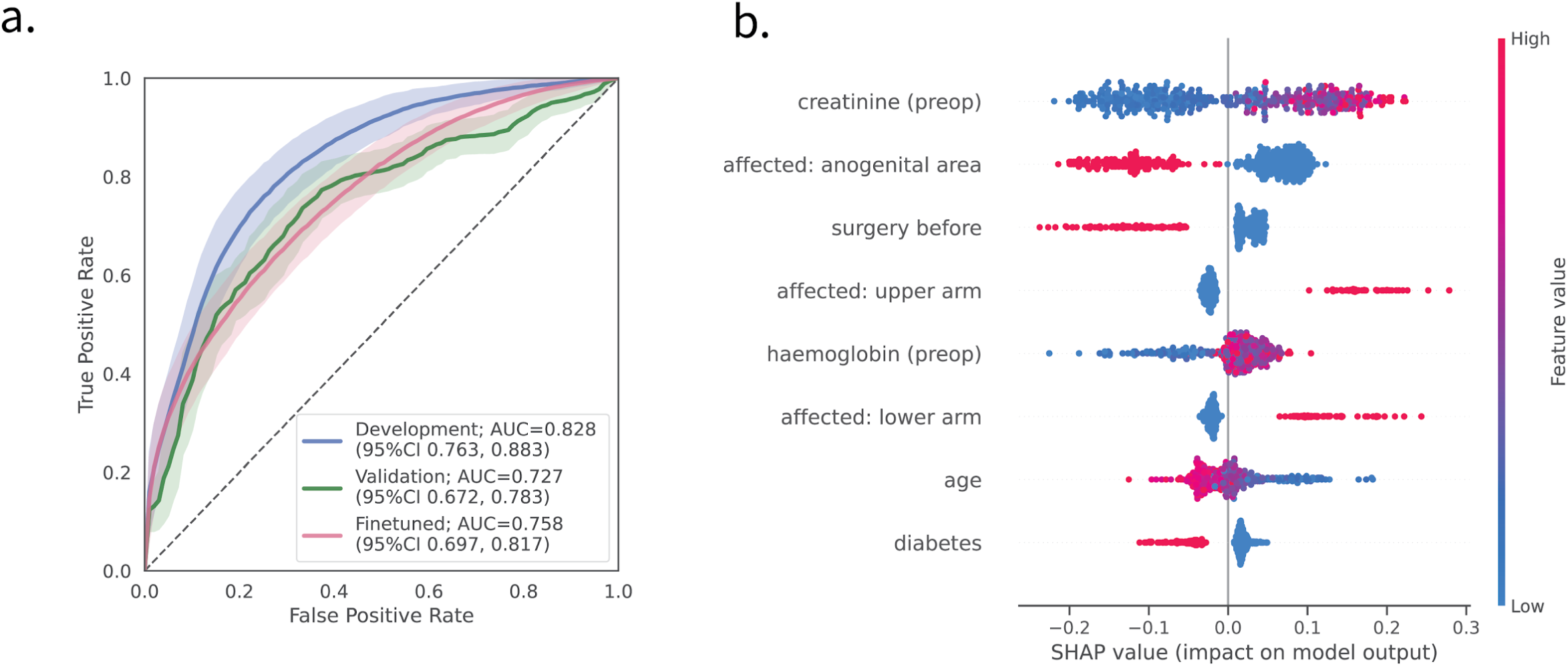
(a) **ROC-curves** comparing the performance of the development (blue), external validation (green), and fine-tuned validation (pink) cohorts. 95%CI: 95% confidence interval derived through 1000 (development) and 10000 (validation) bootstrapped samples (b) **SHAP values for models estimating of GAS aetiology.** Variables are sorted from most impactful (top, creatinine) to least impactful (bottom, diabetes), with every dot representing a patient. Positive SHAP values for a variable indicate a positive contribution to the model’s decision to identify the patient as GAS-positive. Conversely, negative SHAP values indicate a contribution to classifying the patient as GAS-negative. The colour gradient denotes the variable values, with red indicating high values (e.g. age ∼ 70 years) and blue indicating low values (e.g. age ∼ 20 years). SHAP findings are consistent between development and validation cohort.

We sought to validate the performance of the *pre-surgery* model in an external validation cohort, composed of 208 patients with information available on microbial aetiology (208/216 patients, 96.3%). A comparison of the variable characteristics between the development and validation cohorts showed a high degree of similarity (Table 2).

**Table 2.** Comparison of predictive variables’ characteristics between the development and validation cohort. Only variables relevant for the *pre-surgery* model were considered. Detailed variable descriptions can be found in Supplementary Table 3.

|  | **Development cohort**<br>(n=409) | **Validation cohort**<br>(n=208) |
| --- | --- | --- |
| GAS, n (%) | 126 (30.8) | 82 (39.4) |
| age, y, mean (SD) | 58.7 (15.1) | 58.2 (15.5) |
| affected: upper arm, n (%) | 48 (11.7) | 20 (9.3) |
| affected: lower arm, n (%) | 56 (13.7) | 23 (10.7) |
| affected: anogenital area, n (%) | 143 (35.0) | 64 (29.8) |
| surgery before, n (%) | 77 (18.8) | 53 (26.2) |
| diabetes, n (%) | 98 (24.0) | 58 (26.9) |
| creatinine (preop), mean (SD) | 161.2 (121.3) | 153.7 (123.3) |
| haemoglobin (preop), mean (SD) | 11.4 (3.04) | 12.7 (2.5) |

Overall, our model showed a good discriminatory power within the validation cohort with a ROC-AUC of 0.727 (95%CI 0.672, 0.783) (Figure 1a, green). Fine-tuning trained models using 30% of the validation data improved its performance to a ROC-AUC of 0.758 (95%CI 0.696, 0.821) (Figure 1a, pink; Supplementary Table 5). Determination of the optimal threshold by trying to minimise the number of false negatives, yielded an ideal cutoff value of 40%, resulting in a good trade-off between sensitivity and specificity (Table 3).

**Table 3:** Summary of the accuracy of fine-tuned mode at different decision threshold levels. Depicted are the mean and the 95% confidence intervals (in parentheses). Highlighted in bold is the threshold identified as optimal trade-off between model precision and recall when aiming to reduce false-negatives as much as possible. The FPR can be interpreted as the risk of over-treatment, while the FNR indicates the risk for under-treatment. TPR - true positive rate, TNR - true negative rate, FPR - false positive rate, FNR - false negative rate.

| Threshold | Sensitivity<br>(TPR) | Specificity<br>(TNR) | FPR | FNR |
| --- | --- | --- | --- | --- |
| 0 | 1.00 (1.00, 1.00) | 0.00 (0.00, 0.00) | 1.00 (1.00, 1.00) | 0.00 (0.00, 0.00) |
| 0.1 | 0.93 (0.81, 1.00) | 0.29 (0.11, 0.49) | 0.71 (0.51, 0.89) | 0.07 (0.00, 0.19) |
| 0.2 | 0.85 (0.69, 0.97) | 0.46 (0.29, 0.63) | 0.54 (0.37, 0.71) | 0.15 (0.03, 0.31) |
| 0.3 | 0.76 (0.59, 0.90) | 0.59 (0.42, 0.74) | 0.41 (0.26, 0.58) | 0.24 (0.10, 0.41) |
| **0.4** | **0.67 (0.50, 0.82)** | **0.69 (0.55, 0.84)** | **0.31 (0.16, 0.45)** | **0.33 (0.18, 0.50)** |
| 0.5 | 0.57 (0.42, 0.73) | 0.79 (0.64, 0.91) | 0.21 (0.09, 0.36) | 0.43 (0.27, 0.58) |
| 0.6 | 0.46 (0.31, 0.62) | 0.86 (0.73, 0.97) | 0.14 (0.03, 0.27) | 0.54 (0.38, 0.69) |
| 0.7 | 0.36 (0.21, 0.52) | 0.92 (0.80, 0.99) | 0.08 (0.01, 0.20) | 0.64 (0.48, 0.79) |
| 0.8 | 0.25 (0.10, 0.41) | 0.96 (0.87, 1.00) | 0.04 (0.00, 0.13) | 0.75 (0.59, 0.90) |
| 0.9 | 0.13 (0.02, 0.29) | 0.99 (0.95, 1.00) | 0.01 (0.00, 0.05) | 0.87 (0.71, 0.98) |
| 1.0 | 0.00 (0.00, 0.03) | 1.00 (1.00, 1.00) | 0.00 (0.00, 0.00) | 1.00 (0.97, 1.00) |

### Model interpretation

To gain more insight into the decision-making of the model and quantify how much individual variables contributed to predictions, we conducted post-hoc interpretability analysis using SHapley Additive exPlanations (SHAP) (Figure 1b). Inspecting the SHAP values revealed the profound impact of anatomical location on model decisions, with infections occurring in upper extremities being highly predictive of GAS, whereas infections in the anogenital areas hinted at a non-GAS infection. The preoperative creatinine levels, however, appears to be the most influential variable, with values above approximately 110 µmol/L being indicative of GAS (Supplementary Figure 3a).

On the other hand, the presence of diabetes, a surgery conducted four weeks prior to diagnosis, and a higher age (>50 years; Supplementary Figure 3b) were pointed at a low risk for GAS involvement. SHAP findings were consistent between development and validation cohort (Supplementary Figure 4).

### Predicting clinical endpoints: surgical aspects, patient management, and need of organ support

To explore the possibility of estimating clinical endpoints beside the causative microorganisms, we trained predictive models to estimate surgical aspects (risk of amputation, the size of skin defect after fist surgery, the maximal size of skin defect), patient management (length of ICU stay), as well the necessity of organ support (need for RRT within 24 hours after ICU (BL)).

For each outcome, we conducted unsupervised variable selection, yielding between 11 and 16 predictive variables (Supplementary Table 6). Similar to the prediction of GAS aetiology, the performance of several machine-learning models on all time-dependent subsets were compared. Internal validation of the best performing models revealed a lack of predictive performance across all clinical endpoints (Figure 2a, b).

**Figure 2.**
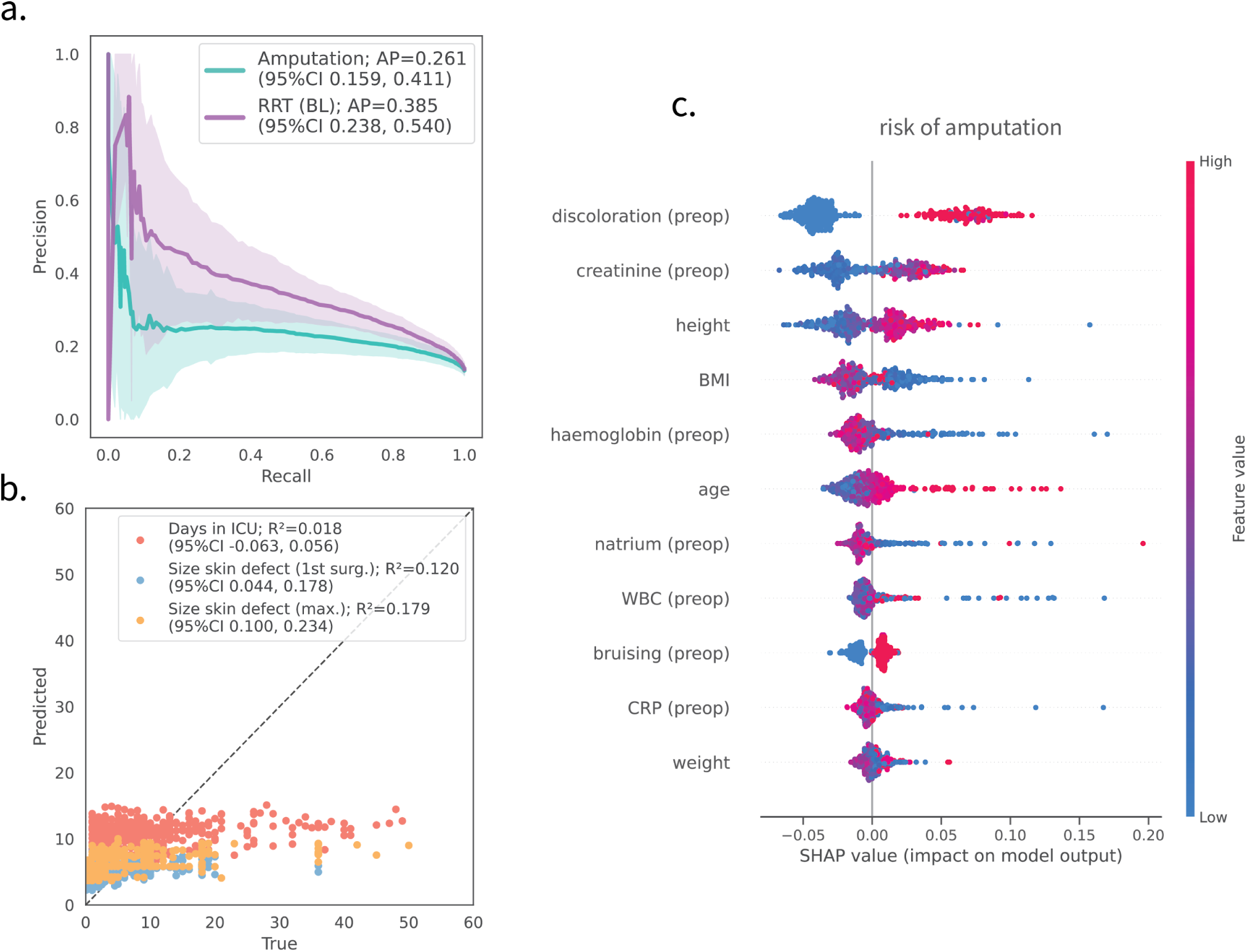
Prediction performance for clinical endpoints revolving around surgical aspects, patient management, and organ support. (a) Precision-recall curves for binary endpoints, including predicting the risk of amputation (turquoise) and need for RRT within the first 24h after ICU admission (baseline, BL). (b) Predicted versus true values for continuous endpoints, including the estimated days in ICU (red, days), size of skin defect after first surgery (blue, percent body surface), and the maximal size of skin defects (orange, percent body surface). (c) SHAP values for models predicting the risk of amputation. Depicted are the mean and the 95% confidence intervals derived through 1000 bootstrapped samples (in parentheses). Summary on included variables and more performance metrics can be found in Supplementary Table 6 and 7 respectively. AP: average precision, R² coefficient of determination

Analysis of SHAP values for surgery-related endpoints, such as the risk of amputation, revealed that models assign high importance to clinically relevant variables, such as the presence of discoloration, creatinine values, of patient’s age (Figure 2c).

## Discussion

In this study we developed and externally validated prediction models for aetiology and various clinical endpoints related to NSTI. Our findings indicate that by using variables available before the first surgery in the referral centre, the presence of GAS aetiology can be successfully predicted, which was confirmed during external validation. Additionally, we showed that fine-tuning trained models on a fraction of the validation data further improved model performances without leading to over-fitting. However, other clinical endpoints explored in this study (i.e. amputation of an extremity, size of the skin defect after surgery, ICU length of Stay, need for Renal Replacement Therapy (RRT)) could not be satisfactorily predicted.

The eight variables relevant to predicting GAS selected through unsupervised variable selection proved to be diverse, ranging from the anatomical location involved to laboratory values. However, all of the selected variables have previously been associated with GAS infections.

Thus, prior investigations in NSTI have revealed a higher occurrence of monomicrobial infections, including GAS, in the upper extremities, while polymicrobial infections were more common in the truncal region (28). This study found a negative association between other surgeries performed last four weeks prior to the NSTI and GAS aetiology. There were 77/409 (19%) patients in the INFECT study with this risk factor (4), whereas numbers were 5/126 (4%) in patients with GAS aetiology (12). The elevated creatinine among GAS NSTI cases reflects that septic shock (65%) and multiorgan dysfunction are common in this group and more frequent than in NSTI of other microbial etiologies (12). Differently, among the GAS NSTI cases there is a negative association to haemoglobin concentrations and, notably, there were no red blood cell transfusions in the entire cohort (4). This may be explained by the higher number of patients with pre-existing comorbidities, more lengthy clinical courses, and more surgery in the preceding four weeks in the non-GAS cases (4,12).

Our assessment of surgical endpoints, such as the risk for amputation and the size of the skin defect, highlights the challenge in objectively evaluating surgical endpoints. Interestingly, we noted that the variables deemed important in predicting the risk of amputation, such as discoloration, bruising, or age, appear to be clinically closely linked to surgical endpoints (29). Despite the logical relevance of these variables, they prove insufficient in predicting skin defect size and whether an amputation of an extremity is performed.

This suggests that either unsupervised variable selection failed to yield objective predictors, and more detailed information about surgical findings upon first debridement would be required. Alternatively, the variability in the decision to perform amputation and the amount of skin that needs to be excised are subjective and influenced by individual surgeons or local guidelines. This latter argumentation is supported by a recent interactive survey on current practice of the debridement of NSTI, which found major variety in the amount of skin that needed to be resected according to Dutch general surgeons and plastic surgeons (30). These results may imply a similar variability in the decision-making process surrounding the need for amputation. A gene predisposition in the STING gene has been linked to amputation and associated with the expression of virulence factors, indicating the complex interaction between host and pathogen influencing NSTI patients’ outcomes (31).

Solely using clinical information ranging from hospital admission to the first 24 hours in ICU, we were unable to accurately estimate the length of stay in the ICU. Given the complexity and heterogeneity of NSTI progression and the fact that patients may need to spend prolonged periods in ICU (4), we believe more longitudinal ICU data is required to predict this outcome successfully.

Previous studies have proven the value of providing decision-support regarding patients’ renal status (32) in predicting the need for renal replacement therapy (RRT). However, the limited number of patients requiring RRT in INFECT resulted in wide confidence intervals, which hindered our ability to draw firm conclusions on the efficacy of our models.

The combination of clinical and computational biology expertise presents a significant strength of this study. By employing a data-driven approach under the guidance of clinical experts, we were able to investigate previously unexplored clinically significant endpoints for NSTI. In addition to utilising the largest NSTI cohort currently available, we conducted rigorous internal validation, as well as externally validated our findings using data from a referral centre not included in the original study. This allowed us to realistically estimate the generalisability of our models. However, the usefulness of external validation to measure a model’s clinical utility and generalisability across different institutions has recently been questioned (26,33). By fine-tuning a pre-trained model with local data, we fully exploited the transferability of machine learning-based models. Our approach showcased the significance of conducting local fine-tuning, enabling the models to adapt to the potential heterogeneity present in data across different timeframes, geographical locations, and facilities, all while avoiding overfitting.

The strengths of our findings should be considered within the context of certain limitations. The development cohort utilised originates from an ICU-focused study, with limited access to pre-hospital data. Due to the uncertainty surrounding the timing of initial symptoms, there exists the possibility of considerable diversity in the progression of the disease among patients, which could potentially affect the performance of our models. Also, we believe the inability to estimate patient management and need of organ support can be partially attributed to the lack of longitudinal data and large imbalances in target labels. Lastly, despite successful external validation, the clinical usefulness of models must still be assessed through prospective validation directly comparing model results with the clinician’s decision.

Our findings demonstrate that prediction of bacterial aetiology is possible, which opens up the possibility of delivering targeted interventions earlier in the disease course of NSTI. The early availability of predictive variables implies that our models can already be used in an emergency room setting. Given the rising incidence of GAS in the Western world (15–17) and studies indicating the beneficial effects of early IVIG administration on survival in GAS patients (12), we believe our findings hold high clinical relevance. Additionally, the conclusions drawn from the prediction of clinical endpoints highlight the need for more research on surgical decision-making.

To the best of our knowledge, this is the first study using machine learning-based methods to estimate aetiology and clinical endpoints relevant to improving NSTI care. Using only eight readily available variables, we developed and validated models capable of estimating the bacterial aetiology prior to surgical debridement. We believe the results of this study to have significant implications for sepsis treatment in patients with NSTI caused by GAS, which may ultimately improve their survival and quality of life.

## Author contributions

SK: Conceptualisation, Methodology, Formal Analysis, Software, Validation, Visualisation, Writing - original draft, review & editing, JS: Funding acquisition, Conceptualisation, Validation, Writing - review & editing, SS: Validation, Funding acquisition, Writing - review & editing, AMV: Funding acquisition, Validation, Writing - review & editing, AP: Funding acquisition, Validation, Writing - review & editing, ANT: Funding acquisition, Project administration, Writing - review & editing, LMPM: Writing - review & editing, JKD: Writing - review & editing, OH: Funding acquisition, Writing - review & editing, EK: Writing - review & editing, MS: Funding acquisition, Project administration, Writing - review & editing, KAM: Validation, Writing - review & editing, VMdS: Supervision, ES: Conceptualization, Methodology, Supervision, Writing - original draft, review & editing

## Transparency declaration

The corresponding author confirms that this manuscript provides a truthful, precise, and clear account of the reported study. All significant aspects of the study have been included. The authors disclose no conflicts of interest.

## Data Availability

All source code and computational results for the models predicting the bacterial GAS aetiology is available at https://github.com/sonjakatz/permit-nsti-gas.

https://github.com/sonjakatz/permit-nsti-gas

## Acknowledgements

The authors are grateful to the members of the INFECT, PerAID and PerMIT projects, for fruitful discussion on the manuscript. This study has received funding from the Swedish Governmental Agency for Innovation Systems (VINNOVA), Innovation Fund Denmark, and the Research Council of Norway under the frame of NordForsk (project No. 90456, PerAID); the Swedish Research Council, Innovation Fund Denmark, the Research Council of Norway, the Netherlands Organisation for Health Research and Development (ZonMW), and DLR Federal Ministry of Education and Research, through the PERMIT project (Personalized Medicine in Infections: from Systems Biomedicine and Immunometabolism to Precision Diagnosis and Stratification Permitting Individualized Therapies, project number 456008002) under the PerMed Joint Transnational call JTC 2018 (Research projects on personalised medicine—smart combination of pre-clinical and clinical research with data and ICT solutions; and from the European Union’s Horizon 2020 research and innovation programme under the Marie Sklodowska-Curie grant agreement No. 860895 TranSYS.

