## Supplementary Material for "A validated model for early prediction of group A streptococcal aetiology and clinical endpoints in necrotising soft tissue infections"

Supplementary Note 1. **Contributors**

### Lead investigators:

#### Coordinating investigator

*Anna Norrby-Teglund*
Centre for Infectious Medicine, Department of Medicine Huddinge
Karolinska Institute, Stockholm, Sweden

#### National site investigators

**Copenhagen**
*Ole Hyldegaard*
Department of Anaesthesia, Centre of Head and Orthopaedics
Copenhagen University Hospital, Rigshospitalet, Copenhagen, Denmark

**Stockholm**
*Michael Nekludov*
Department of Anaesthesia, Surgical Services and Intensive Care
Karolinska Institute, Karolinska University Hospital, Stockholm, Sweden

**Karlskrona**
*Ylva Karlsson*
Department of Anaesthesia and Intensive Care
Blekinge County Hospital, Karlskrona, Sweden

**Gothenburg**
*Per Arnell*
Department of Anaesthesia and Intensive Care
Sahlgrenska University Hospital, Gothenburg, Sweden

**Bergen**
*Steinar Skrede*
Department of Medicine
Haukeland University Hospital, Bergen, Norway
Department of Clinical Science,
University of Bergen, Bergen, Norway

### The INFECT study group:

**Karolinska Institutet, Karolinska University Hospital, Stockholm Sweden**
Anna Norrby-Teglund (Co-ordinator), Mattias Svensson (Project Manager), Muhammad Afzal, Helena Bergsten, Lydia Bosnak, Bavya Chakrakodi, Puran Chen, Johanna Emgård, Linda Johansson, Julius Juarez, Srikanth Mairpady Shambat, Nikola Siemens, Johanna Snäll, Julia Uhlman, Takeaki Wajima.

**Copenhagen University Hospital, Rigshospitalet, Copenhagen, Denmark**
Ole Hyldegaard (Team Leader, Co-ordinator clinical partners), Martin B. Madsen (Clinical Database), Daniel Bidstrup, Nina F. Bærnthsen, Julie V. Clausen, Anna Damgaard, Gladis H. Frendø, Martin Forchammer, Marco Hansen, Morten FF Hedetoft, Karen L. Hilsted, Diana Isaksen, Erik C. Jansen, Josefine Kofoed, Anette Lilja, Lærke B. Madsen, Rasmus Müller, Isabel S. Nielsen, Emilie MJ Pedersen, Marie W. Petersen, Anders Perner, Peter V. Polzik, Frederikke Ravn

**Karolinska University Hospital, Stockholm, Sweden**
Michael Nekludov (Team Leader), Folke Lind, Anders Kjellberg, Erik von Oelreich, Peter Kronlund, Sverre Kullberg, Ola Friman, Lisa Hellgren, Anna Granström, Anna Schenning, Sandra Carlsson

**Blekingesjukhuset Karlskrona**
Ylva Karlsson (Team Leader), Dag Benoni

**Sahlgrenska University Hospital Ostra, Gothenburg, Sweden**
Per Arnell (Team Leader), Hans Lycke, Joakim Trogen, Kerstin Ohlauson University of Bergen, Haukeland University Hospital, Bergen, Norway Steinar Skrede (Team Leader), Trond Bruun, Torbjørn Nedrebø, Oddvar Oppegaard, Eivind Rath, Marianne Sævik, Hanne Søyland

**Helmholtz Center for Infection Research, Braunschweig, Germany**
Dietmar H. Pieper (Team Leader), Singh Chhatwal, Andreas Itzek, Anshu Babbar, Robert Thänert, Jörn Hoßmann, Eva Medina, Domenica Hamisch, Israel Barrantes, Patric Nitsche-Schmitz, Astrid Dröge, Katja Mummenbrauer.

**Wageningen University and Research, Wageningen, The Netherlands**
Vitor Martins Dos Santos (Team Leader), Edoardo Saccenti, Jasper Koehorst, Peter Schaap

**Université Lyon 1, Lyon, France**
Francois Vandenesh (Team Leader), Sylvere Bastien, Jessica Baude, Anne Tristan.

**LifeGlimmer GmBH, Berlin, Germany**
Vitor Martin dos Santos (Team Leader), Erno Lindfors, Francois Bergey

**Cube Dx GmbH, Sankt Valentin, Austria**
Christoph Reschreiter (team Leader), Bernhard Ronacher， Matthias Pilecky

**Tel Aviv University, Tel Aviv, Israel.**
Eytan Ruppin (Team Leader), Matthew Oberhardt, Raphy Zarecky.

**University of North Dakota, Grand Forks, USA**
Malak Kotb (Team Leader), Karthickeyan Chellakrishnan, Santhosh Mukundan, Suba Nokala,

**The Lee Spark NF Foundation, UK**
Doreen Marsden (Team Leader).

### PerAID/PerMIT Study Group - non-authors involved

Mattias Svensson[1], Kristoffer Strålin[1,2], Trond Bruun[3,4], Oddvar Oppegaard[3,4], Knut Anders Mosevoll[3,4], Jan Kristian Damås[5,6,7], P.P.M van Zuijlen[8,9,10], Marco Anteghini[11,12]

1 Center for Infectious Medicine, Department of Medicine Huddinge, Karolinska Institute, Stockholm, Sweden
2 Department of Medicine, Huddinge, Karolinska Institutet, Stockholm, SWEDEN
3 Department of Medicine, Haukeland University Hospital, Bergen, Norway
4 Department of Clinical Science, University of Bergen, Bergen, Norway
5 Gemini Center for Sepsis Research, Department of Circulation and Medical Imaging, NTNU Norwegian University of Science and Technology, Trondheim, Norway
6 Centre of Molecular Inflammation Research, Department of Clinical and Molecular Medicine, NTNU, Norwegian University of Science and Technology, Trondheim, Norway
7 Department of Infectious Diseases, St Olavs Hospital, Trondheim University Hospital, Trondheim, Norway
8 Burn Centre, Department of Plastic, Reconstructive and Hand Surgery, Red Cross Hospital, Beverwijk, The Netherlands
9 Pediatric Surgical Centre, Emma Children's Hospital, Amsterdam UMC, Amsterdam, The Netherlands
10 Department of Plastic, Reconstructive and Hand Surgery, Amsterdam Movement Sciences Amsterdam UMC, Amsterdam, The Netherlands
11 LifeGlimmer GmbH, Berlin, Germany
12 Laboratory of Systems and Synthetic Biology, Wageningen University and Research, Wageningen, The Netherlands

Supplementary Note 2. **Overview of hyperparameters tuned**

**Classification algorithms:**

Logistic Regression (LR):

- *penalty*: none, l2
- *C*: 0.001,0.01,0.1,1,10,100,1000

Gaussian process classifier (GPC):

- *kernel*: DotProduct, Matern, RationalQuadratic, WhiteKernel

Gaussian Naive Bayes (GNB):

- *var_smoothing*: 100 steps from -9 until +9

Random Forest Classifier (RFC):

- *n_estimators*: 100, 300, 700
- *max_depth*: 2,4,6
- *max_features*: 2,4,6

**Regression algorithms:**

Lasso regression:

- *selection*: “random”
- *alpha*: 1.0, 0.7, 0.5

Ridge regression

- *alpha*: 1.0, 0.7, 0.5

Elastic nets:

- *l1_ratio*=0.5
- *selection*="random"
- *alpha*: 1.0, 0.7, 0.5

Multi-layer perceptron regression:

- *activation*: "relu"
- *batch_size*: 64
- *early_stopping*: True
- *solver*: “adam”
- *alpha*: 1, 0.1, 0.01, 0.001, 0.0001
- *hidden_layer_size*: (10, 3), (11, 3), (8, 3), (7, 3), (9, 3)

Random Forest Regressor:

- *n_estimators*: 100, 300
- *max_depth*: 2,4
- *max_features*: 2,4

Supplementary Figure 1. **Structured flowcharts detailing the (a) developmental and validation process of the machine learning models (b) time-dependent data dissection.** (a) This conceptual pipeline was systematically implemented for every clinical outcome assessed. External validation was exclusively pursued concerning the bacterial etiology (presence of Group A Streptococcus).(b) The INFECT study cohort was split into the time-dependent subsets *entry, pre-* and *post-surgery* at referral center, *baseline*, depending on the clinical availability of the data. n: number of patients/iterations; LOOCV: leave-one-out cross-validation; LR: logistic regression, GPC: Gaussian process classifier, GNB: Gaussian naive bayes; RFC: Random Forest classifier; Lasso: Lasso regression, Ridge: Ridge regression, EN: elastic net regression, GPR: Gaussian process regression, MLP: multi-layer perceptron; RFR: Random Forest regression.

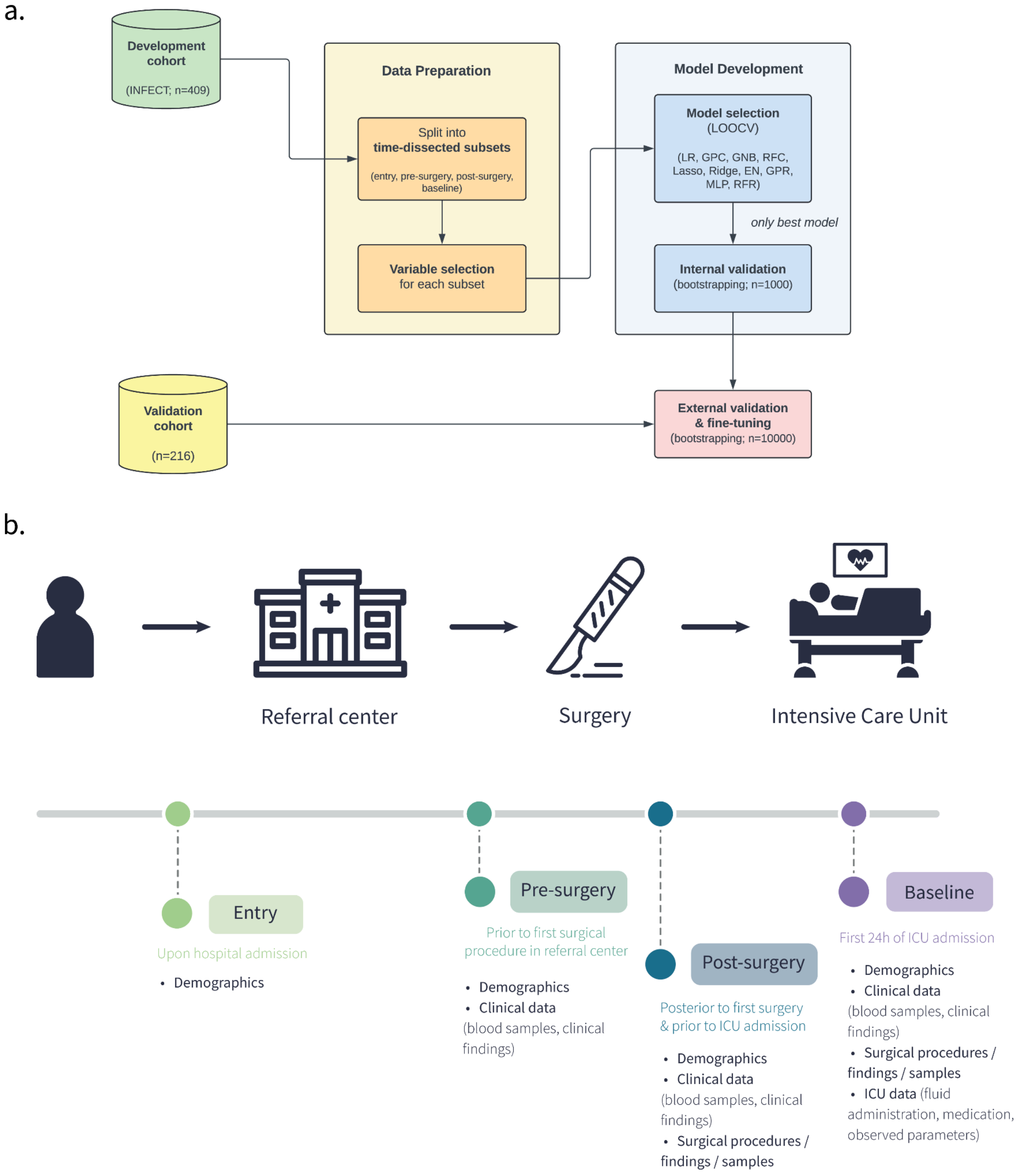

Supplementary Figure 2. **Comparison performance between different time-dependent datasets for the estimation of the presence of GAS.** (left) ROC curve (right) Precision-Recall curve. ENTRY (upon hospital admission), PRESURGERY (prior to first surgical procedure), POSTSURGERY (posterior to first surgical procedure and prior to ICU admission), BL (baseline; first 24 h of ICU admission). AUC: Area under the ROC curve, AP: average precision.

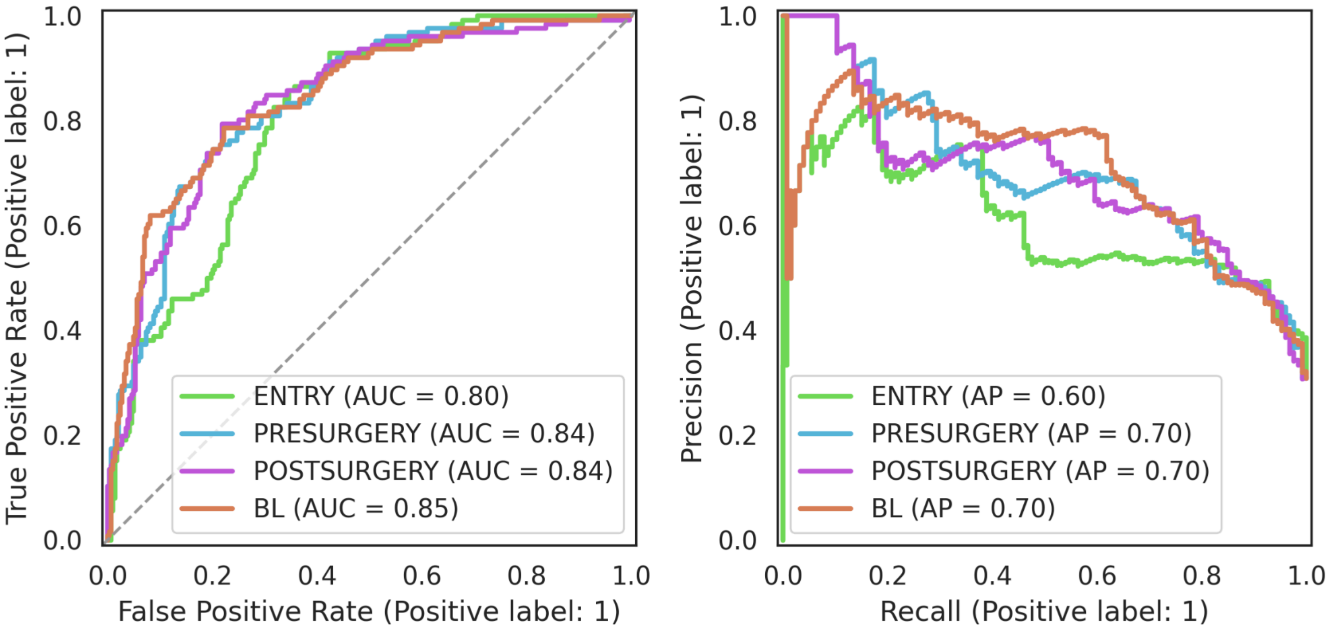

Supplementary Figure 3. **SHAP dependence plots** for (a) age (b) creatinine (c) haemoglobin. The colour gradient denotes the variable values, with red indicating high values (e.g. age ~ 70 years) and blue indicating low values  (e.g. age ~ 20 years).

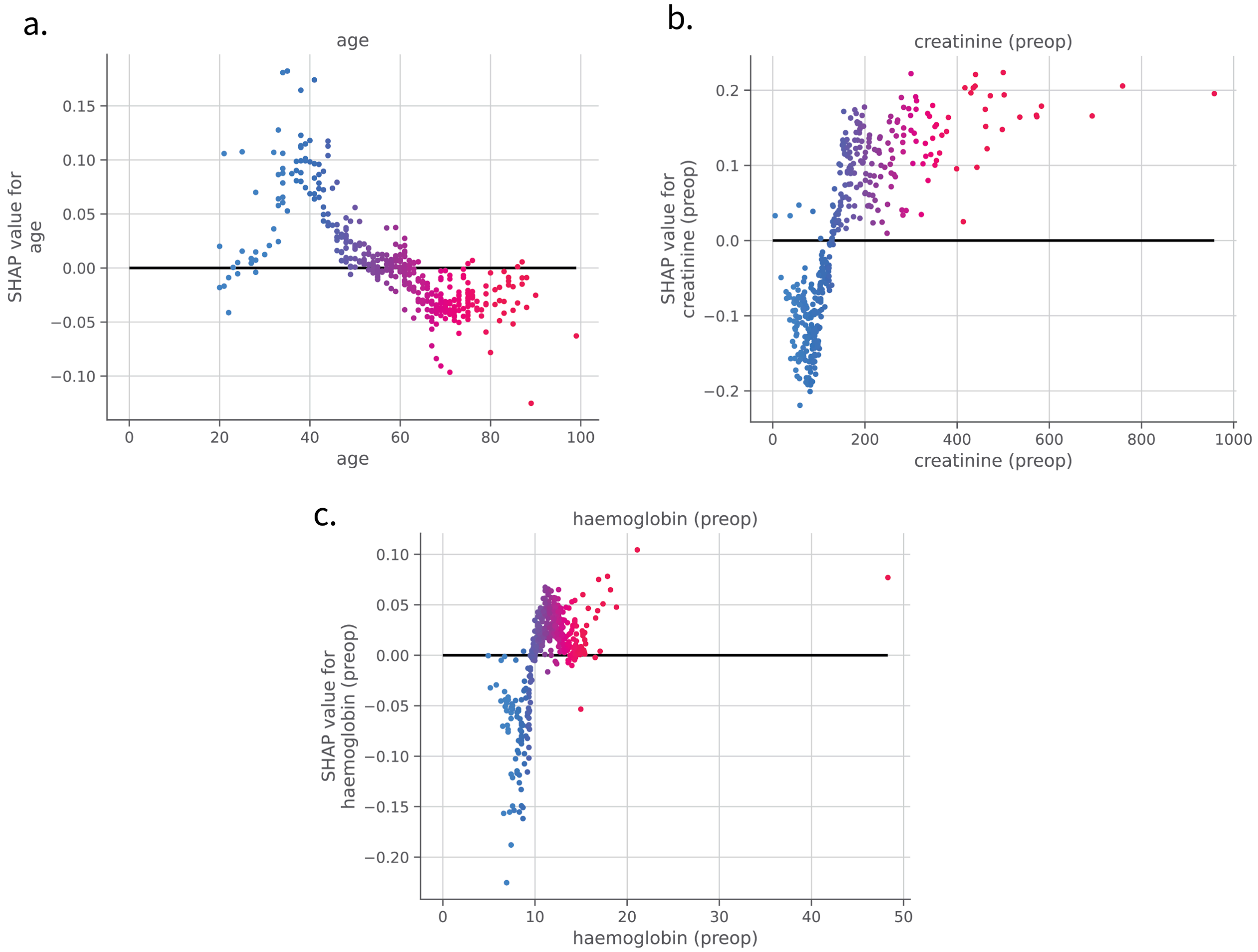

Supplementary Figure 4. **SHAP values for models estimating the presence of GAS with the external validation cohort.** Variables are sorted from most impactful (top, creatinine) to least impactful (bottom, diabetes), with every dot representing a patient. Positive SHAP values for a variable indicate a positive contribution to the model's decision to identify the patient as GAS-positive. Conversely, negative SHAP values indicate a contribution to classifying the patient as GAS-negative. The colour gradient denotes the variable values, with red indicating high values (e.g. age ~ 70 years) and blue indicating low values (e.g. age ~ 20 years).

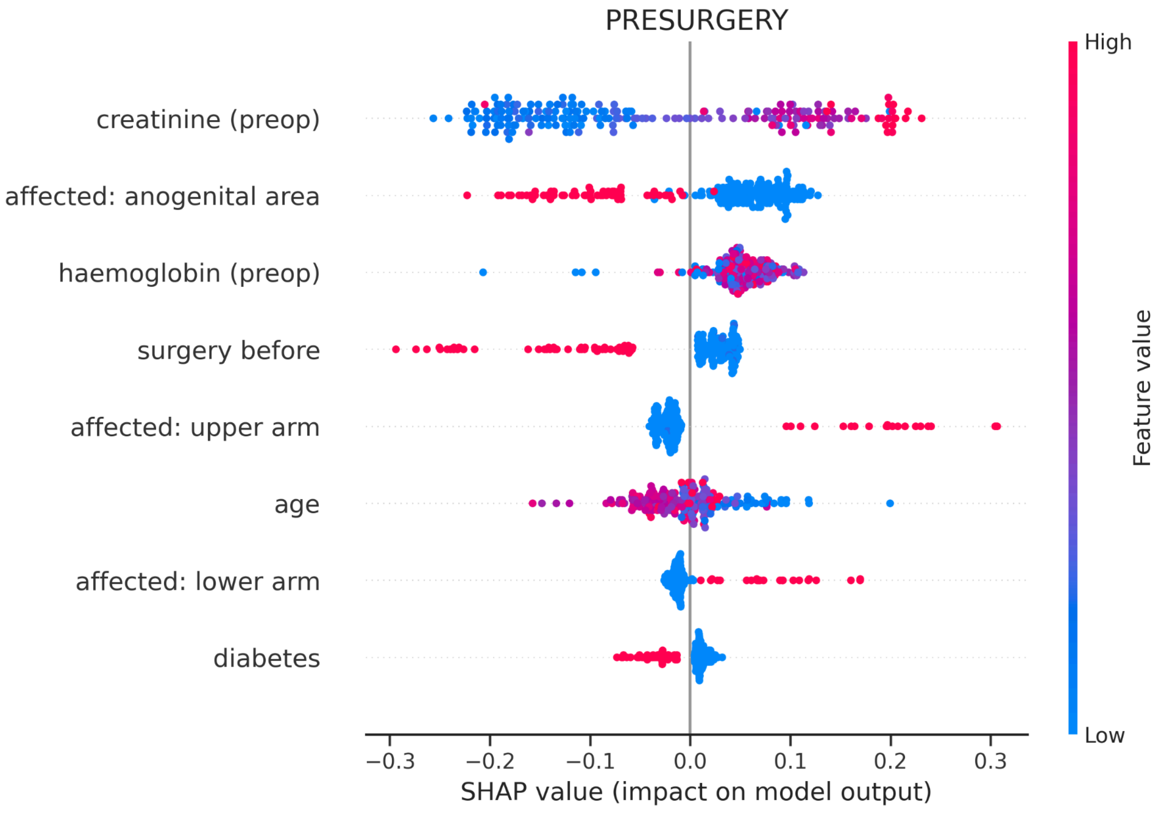

Supplementary Table 1: **Overview for the number of patients included and data subsets assessed for each clinical outcome**.

| **Outcome category** | **Outcome** | **No. of patients (n)** | **Time-dependent data subsets** | **Prediction task** | **No. of patients (%) or mean ± SD** |
|---|---|---|---|---|---|
| causative microbes | Presence of GAS | 409 | Entry, pre-surgery, post-surgery, baseline | classification (y/n) | 126 (30.8%) |
| surgical aspects | Risk of amputation | 409 | Entry, pre-surgery | classification (y/n) | 54 (13.2%) |
| | Size of skin defect (after first surgery) | 391 | Entry, pre-surgery | regression (pct. of body surface) | 4.9 ± 5.2 |
| | Size of skin defect (maximal) | 409 | Post-surgery, baseline | regression (pct. of body surface) | 6.4 ± 7.3 |
| patient management | Length of ICU stay | 402 | Entry, pre-surgery, post-surgery, baseline | regression (days) | 10.7 ± 10.8 |
| organ support | Need for RRT (within 24h after ICU admission) | 409 | Entry, pre-surgery, post-surgery | classification (y/n) | 57 (13.9%) |

Supplementary Table 2: Summary of number of patients and variables in each time-dependent data subset. Details on the timeline indicating the timing of each categorised clinical variable collected can be found in Supplementary Figure 2.

| | **No. of patients (n)** | **No. of variables (n)** |
|---|---|---|
| **Entry** | 409 | 45 |
| **Pre-surgery** | 409 | 56 |
| **Post-surgery** | 409 | 723 |
| **Baseline** | 409 | 762 |

Supplementary Table 3: **Detailed description for variables used in the estimation of the presence of GAS.**

| Variable name | Description |
|---|---|
| age | age at admission [years] |
| affected: upper arm | affection of upper arm[1] [y/n] |
| affected: lower arm | affection of lower arm[1] [y/n] |
| affected: anogenital area | affection of anogenital area[1] [y/n] |
| surgery before | surgery within 4 weeks previous of NSTI [y/n] |
| diabetes | diabetes [y/n] |
| creatinine (preop) | Highest preoperative[2] creatinine [μmol/L] |
| haemoglobin (preop) | Lowest preoperative[2] haemoglobin [mmol/L] |
| creatinine (preadmission) | Highest preadmission[3] creatinine [μmol/L] |
| Lowest systolic BP (BL) | Lowest BL[4] systolic blood pressure [mmHg] |
| Creatinine (BL) | Highest BL[4] creatinine [μmol/L] |
| Noradrenaline (BL) | Highest noradrenaline infusion rate at BL[4] [μg/kg/min] |
| Platelets (BL) | Lowest BL[4] platelets [$10^9$/L] |
| Lactate (BL) | Highest BL[4] lactate level [mmol/L] |
| Glucose (BL) | Highest BL[4] glucose [mmol/L] |
| Anatomical site sampled | Anatomical site[5] specimen was sampled from |

[1] at arrival at sepcialized hospital

[2] before the first surgery, which is before ICU admission

[3] upon ICU admission

[4] during the first 24 hours in the ICU

[5] one of the following: head/neck, u. arm, l. arm, hand, finger, thorax, abdomen, ano-gen., u. leg, l. leg, foot, toe

Supplementary Table 4. **Performance comparison of several machine learning-based classifiers for the prediction of GAS aetiology for each time-dissected data subset.** Displayed are the mean performances across leave-one-out cross-validation (LOOCV). log: Logistic Regression, gpc: Gaussian Process Classifier, rfc: Random Forest Classifier, Acc.: balanced accuracy, Prec.: precision, Brier: Brier score, ROC AUC: area under the receiver-operator curve, Ave. prec.: average precision

| Entry | | | | | | |
|---|---|---|---|---|---|---|
| **Model** | **Prec.** | **Recall** | **F1-score** | **Acc.** | **ROC AUC** | **Ave. prec.** |
| log | 0.520 | 0.825 | 0.638 | 0.743 | 0.792 | 0.595 |
| gpc | 0.517 | 0.825 | 0.636 | 0.741 | 0.794 | 0.608 |
| rfc | 0.524 | 0.857 | 0.651 | 0.755 | 0.804 | 0.604 |
| **Pre-surgery** | | | | | | |
| **Model** | **Prec.** | **Recall** | **F1-score** | **Acc.** | **ROC AUC** | **Ave. prec.** |
| log | 0.582 | 0.762 | 0.660 | 0.759 | 0.825 | 0.659 |
| gpc | 0.551 | 0.810 | 0.656 | 0.758 | 0.827 | 0.646 |
| rfc | 0.633 | 0.738 | 0.681 | 0.774 | 0.838 | 0.704 |
| **Post-surgery** | | | | | | |
| **Model** | **Prec.** | **Recall** | **F1-score** | **Acc.** | **ROC AUC** | **Ave. prec.** |
| log | 0.643 | 0.714 | 0.677 | 0.769 | 0.840 | 0.674 |
| gpc | 0.652 | 0.730 | 0.689 | 0.779 | 0.850 | 0.693 |
| rfc | 0.617 | 0.794 | 0.694 | 0.787 | 0.839 | 0.700 |
| **Baseline** | | | | | | |
| **Model** | **Prec.** | **Recall** | **F1-score** | **Acc.** | **ROC AUC** | **Ave. prec.** |
| log | 0.593 | 0.810 | 0.685 | 0.781 | 0.844 | 0.660 |
| gpc | 0.674 | 0.722 | 0.697 | 0.783 | 0.860 | 0.704 |
| rfc | 0.757 | 0.619 | 0.681 | 0.765 | 0.846 | 0.704 |

Supplementary Table 5. **Model performances in estimating GAS involvement for time-dissected datasets.** Depicted are the mean and the 95% confidence intervals (in parentheses). Acc.: balanced accuracy, Prec.: precision, Brier: Brier score, ROC AUC: area under the receiver-operator curve, Ave. prec.: average precision

| | **Entry** | **Pre-surgery** | **Post-surgery** | **Baseline** | **Pre-surgery (fine-tuned)** |
|---|---|---|---|---|---|
| **Acc.** | 0.652 (0.574,0.735) | 0.726 (0.642,0.796) | 0.723 (0.644,0.791) | 0.727 (0.658,0.799) | 0.677 (0.617, 0.737) |
| **Prec.** | 0.572 (0.435,0.724) | 0.666 (0.548,0.784) | 0.683 (0.556,0.811) | 0.729 (0.600,0.867) | 0.644 (0.534, 0.776) |
| **Recall** | 0.463 (0.267,0.698) | 0.583 (0.391,0.739) | 0.564 (0.396,0.720) | 0.547 (0.396,0.700) | 0.568 (0.417, 0.726) |
| **F1-score** | 0.502 (0.351,0.632) | 0.617 (0.483,0.719) | 0.613 (0.486,0.714) | 0.621 (0.500,0.729) | 0.598 (0.509, 0.682) |
| **Brier** | 0.170 (0.142,0.203) | 0.152 (0.128,0.183) | 0.149 (0.125,0.180) | 0.147 (0.125,0.172) | 0.199 (0.168, 0.238) |
| **ROC AUC** | 0.794 (0.733,0.851) | 0.828 (0.763,0.883) | 0.836 (0.775,0.891) | 0.839 (0.779,0.894) | 0.758 (0.697, 0.817) |
| **Ave. prec** | 0.617 (0.494,0.719) | 0.684 (0.568,0.787) | 0.685 (0.570,0.788) | 0.703 (0.592,0.807) | 0.701 (0.604, 0.780) |

Supplementary Table 6. **Overview of variables yielded through unsupervised variable selection** for prediction of surgical, patient management, and organ support outcomes. BMI: Body Mass Index, BL: baseline (24 hours after ICu admission), CRP: c-reactive protein, preop: preoperative, KDIGO: Kidney Disease Improving Global Outcomes, WBC: white blood cell count,

| **Risk of amputation** | **Size of skin defect (after first surgery)** | **Size of skin defect (maximal)** | **Days spent in ICU** | **Need for RRT (24h after ICU admission)** | **Need for RRT (90 days)** |
|---|---|---|---|---|---|
| Weight | Weight | BP (highest BL) | No. of blood samples taken | Weight | Pulse (lowest BL) |
| Height | Height | Bilirubin (highest BL) | Discoloration (preop) | Height | Carbamid (highest BL) |
| Discoloration (preop) | Bullae (preop) | No. of blood samples (1st surgery) | WBC (preop) | Discoloration (preop) | Kalium (highest BL) |
| Bruising (preop) | WBC (preop) | Blood collection mode | CRP (preop) | WBC (preop) | Bicarbonate (lowest BL) |
| WBC (preop) | CRP (preop) | Bullae (preop) | creatinine (preop) | CRP (preop) | Urine output (BL) |
| CRP (preop) | creatinine (preop) | WBC (preop) | Haemoglobin (preop) | creatinine (preop) | creatinine (preop) |
| creatinine (preop) | Natrium (preop) | Blood products (baseline) | Age | Natrium (preop) | creatinine (BL) |
| Natrium (preop) | Haemoglobin (preop) | Center code | affected: head/neck | Haemoglobin (preop) | Noradrenaline (max BL) |
| Haemoglobin (preop) | BMI | Bullae (during 1st surgery) | Varicella | BMI | pH (lowest BL) |
| BMI | Center code | Haemoglobin (preop) | Size of skin defect (after first surgery) | Age | SBE (lowest BL) |
| Age | Age | CRP (preop) | | affected: upper leg | Lactate (highest BL) |
| | affected: abdomen | | | Cardiovascular disease present | Crystalloids (BL) |
| | affected: upper leg | | | | Accumulated fluids (BL) |
| | affected: lower leg | | | | KDIGO stage |
| | | | | | creatinine (preadmission) |
| | | | | | WBC (preop) |
| **n = 11** | **n = 14** | **n = 11** | **n = 10** | **n = 12** | **n = 16** |

Supplementary Table 7. **Prediction performance for clinical endpoints revolving around surgical aspects, patient management, and organ support.** Ave. prec. : average precision, $R^2$ coefficient of determination, MAE: mean absolute error, MSE: mean squared error

| | **Risk of amputation** | **Size of skin defect (after first surgery)** | **Size of skin defect (maximal)** | **Days spent in ICU** | **Need for RRT (24h after ICU admission)** |
|---|---|---|---|---|---|
| **Acc.** | 0.503 (0.483, 0.546) | - | - | - | 0.543 (0.493, 0.610) |
| **Prec.** | 0.137 (0.000, 1.000) | - | - | - | 0.467 (0.000, 1.000) |
| **Recall** | 0.020 (0.000, 0.106) | - | - | - | 0.109 (0.000, 0.250) |
| **F1-score** | 0.033 (0.000, 0.182) | - | - | - | 0.168 (0.000, 0.345) |
| **Brier** | 0.113 (0.092, 0.136) | - | - | - | 0.105 (0.083, 0.125) |
| **Ave. prec.** | 0.261 (0.159, 0.411) | - | - | - | 0.385 (0.238, 0.540) |
| **$R^2$** | - | 0.120 (0.044, 0.178) | 0.179 (0.100,0.234) | 0.018 (-0.063,0.056) | - |
| **MAE** | - | 3.500 (3.144, 3.831) | 4.383 (3.905, 4.879) | 7.047 (6.194,7.976) | - |
| **MSE** | - | 24.086 (16.334, 34.611) | 43.328 (27.789, 66.054) | 111.952 (65.077, 172.308) | - |
